# Association of *HOXB13* G84E with prostate cancer among 592,158 men

**DOI:** 10.1101/2024.10.15.24315450

**Authors:** Taylor B. Crawford, Tyler Nelson, Roshan Karunamuni, Heena Desai, Ryan Hausler, Craig Teerlink, Hannah Carter, Meghana S. Pagadala, Patrick R. Alba, Scott L. DuVall, Morgan E. Danowski, Charles A. Brunette, Dmitry Ratner, Isla P. Garraway, Brent S. Rose, Kathleen A. Cooney, Jason L. Vassy, Richard L. Hauger, Julie A. Lynch, Tyler M. Seibert, Kara N. Maxwell

## Abstract

The *HOXB13* c.G251G>A:p.G84E (rs138213197) variant confers an increased risk of prostate cancer (PCa). Minimal data exist from population-based cohorts to guide genetic counseling. We identified men heterozygous for *HOXB13* p.G84E or homozygous wild-type among 592,158 male participants in the Million Veteran Program (MVP). 1660 (0.3%) men in MVP were heterozygous for *HOXB13* p.G84E. Results from Cox proportional hazards models demonstrated significant associations between *HOXB13* p.G84E and risk of any PCa (HR 3.23, 95%CI 2.97-3.53, p<2.0E-16), metastatic PCa (HR 2.96, 95%CI 2.30-3.81, p<2.0E-16), and PCa death (HR 2.65, 95%CI 1.67-4.21, p=3.8E-05). In a subset of MVP participants who underwent prostate biopsy at VA (n=36,321), a multivariable logistic regression model controlling for known PCa risk factors showed that *HOXB13* p.G84E heterozygotes had a higher risk of PCa diagnosis (OR 2.60, 95%CI 1.94-3.52, p<0.001). *HOXB13* p.G84E heterozygotes had slightly younger age of PCa diagnosis but similar distribution of Gleason scores and rates of de novo, any, and castrate resistant metastatic disease. In the largest cohort of men with the *HOXB13* p.G84E variant studied to date, we show this variant confers a moderately increased lifetime risk of PCa. Further work is needed to determine if early PCa screening in *HOXB13* p.G84E heterozygotes improves outcomes.

**Patient Summary:** Genetic testing may reveal a variant in *HOXB13* called p.G84E. This variant increases a person’s risk of prostate cancer by three-fold. However, the prostate cancer that develops is not more aggressive, although it may occur at younger ages.

## Main

Prostate cancer (PCa) is the second leading cause of cancer death in males^1^. Identifying males at highest risk of aggressive PCa may help focus early detection programs on patients most likely to benefit from screening and reduce the burden of overdiagnosis and unnecessary procedures^2^.

The *HOXB13* c.G251G>A;p.G84E (rs138213197) pathogenic germline variant was identified as an autosomal dominant PCa risk allele in individuals with a strong family history of PCa^3^. The variant occurs on a common haplotype consistent with a founder effect and is most commonly observed in White men of European descent^4^. Subsequent studies in high-risk (*i*.*e*. strong family history) cohorts found that *HOXB13* p.G84E conferred an 3-5-fold increased risk of PCa^4–6^; however, these studies may have overestimated risk. Further, whether *HOXB13* p.G84E confers risk of more aggressive PCa remains controversial^5–7^.

Herein, we report age-specific risk estimates for the development of any, metastatic, and fatal PCa in *HOXB13* p.G84E heterozygotes among 592,158 male participants in the Million Veteran Program (MVP), a diverse healthcare-system-based genotyped cohort. We additionally report clinical and pathological characteristics of PCa in the largest reported cohort of male individuals heterozygous for *HOXB13* p.G84E (n=1,660).

We obtained genetic data from the MVP Genisis^8^ and identified *HOXB13* p.G84E heterozygotes and wild-type controls (**Supplemental Methods**). Heterozygote frequencies in MVP and gnomADv4 were not statistically significantly different, demonstrating high quality genotyping at this locus (**Table S1**). Clinical and pathological data were obtained from the VA Corporate Data Warehouse (CDW) and the Prostate Cancer Data Core for all individuals who underwent first prostate biopsy within the VA^9^.

We assessed for association of *HOXB13* p.G84E with age at any PCa diagnosis, metastatic PCa, and death from PCa using Cox proportional hazards models^10^ and clinical/pathological variables using chi-squared (binary variables) and Kruskal-Wallis (continuous variables) tests in the 592,158 men genotyped in MVP. In a subset of patients undergoing their first prostate biopsy in VA, associations between *HOXB13* p.G84E status and risk of any PCa on first biopsy and aggressive PCa were determined using multivariable logistic regression models^9^.

Among 592,158 men in MVP, 1,660 (0.28%) were heterozygous for the *HOXB13* c.G251G>A:p.G84E (rs138213197) variant, the majority (94.5%) of whom identified as White, compared to 71.8% of the full cohort (**Table S2**). Among *HOXB13* p.G84E heterozygotes, 519 (31.3%) had a diagnosis of PCa compared to 71,500 (12.1%) homozygous wild-type individuals (p<0.001) (**Table 1**). In a Cox proportional hazards model, *HOXB13* p.G84E heterozygotes had a significantly higher risk of any PCa (HR 3.23, 95% CI 2.97-3.53, p<2.0E-16), metastatic PCa (HR 2.96, 95%CI 2.30-3.81, p<2.0E-16), and PCa death (HR 2.65, 95% CI 1.67-4.21, p=3.8E- 05) (**Figure 1A-C**).

**Table 1:**
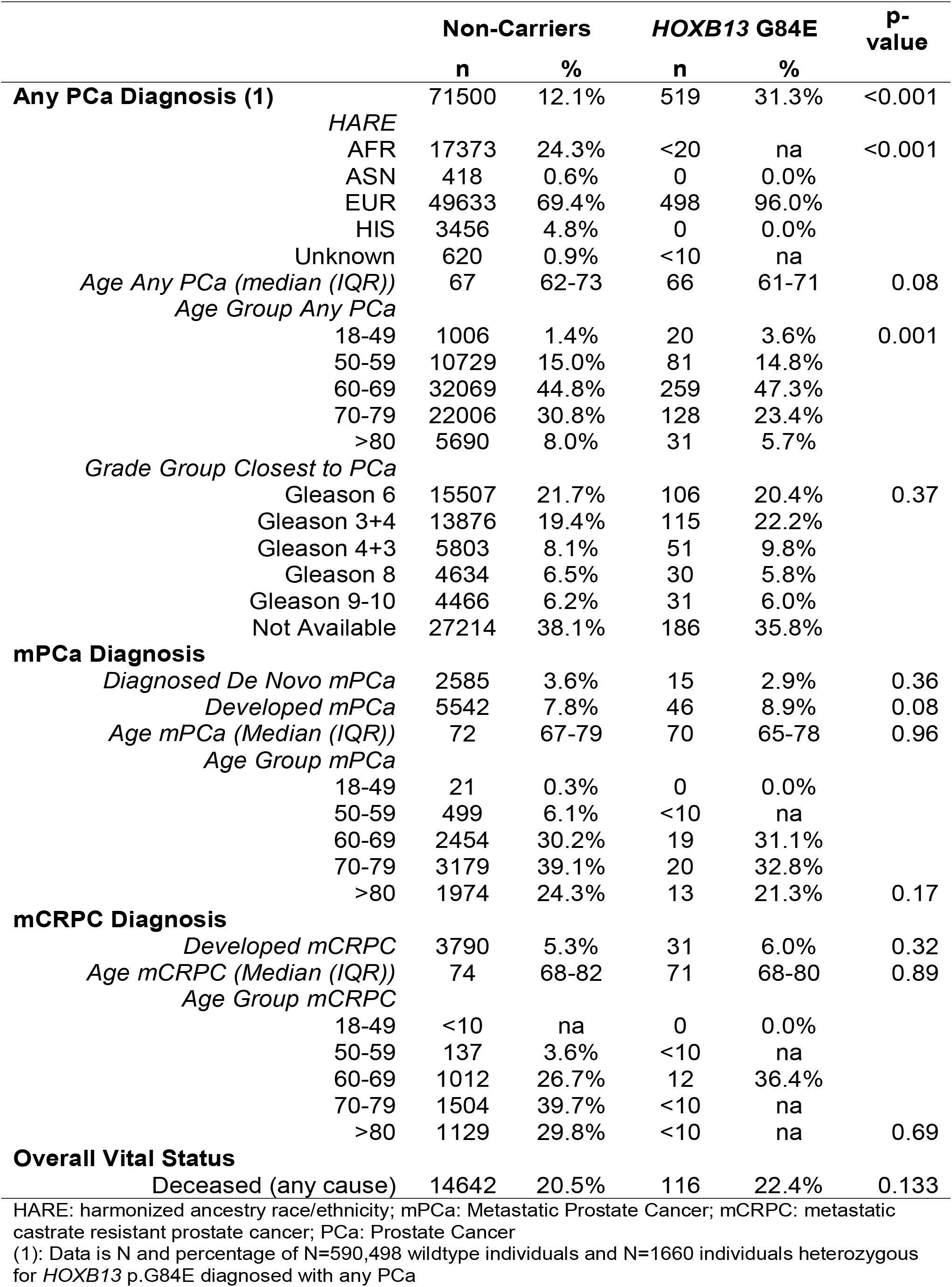
Clinicopathological characteristics of PCa diagnosed in individuals heterozygous for *HOXB13* p.G84E.

**Figure 1:**
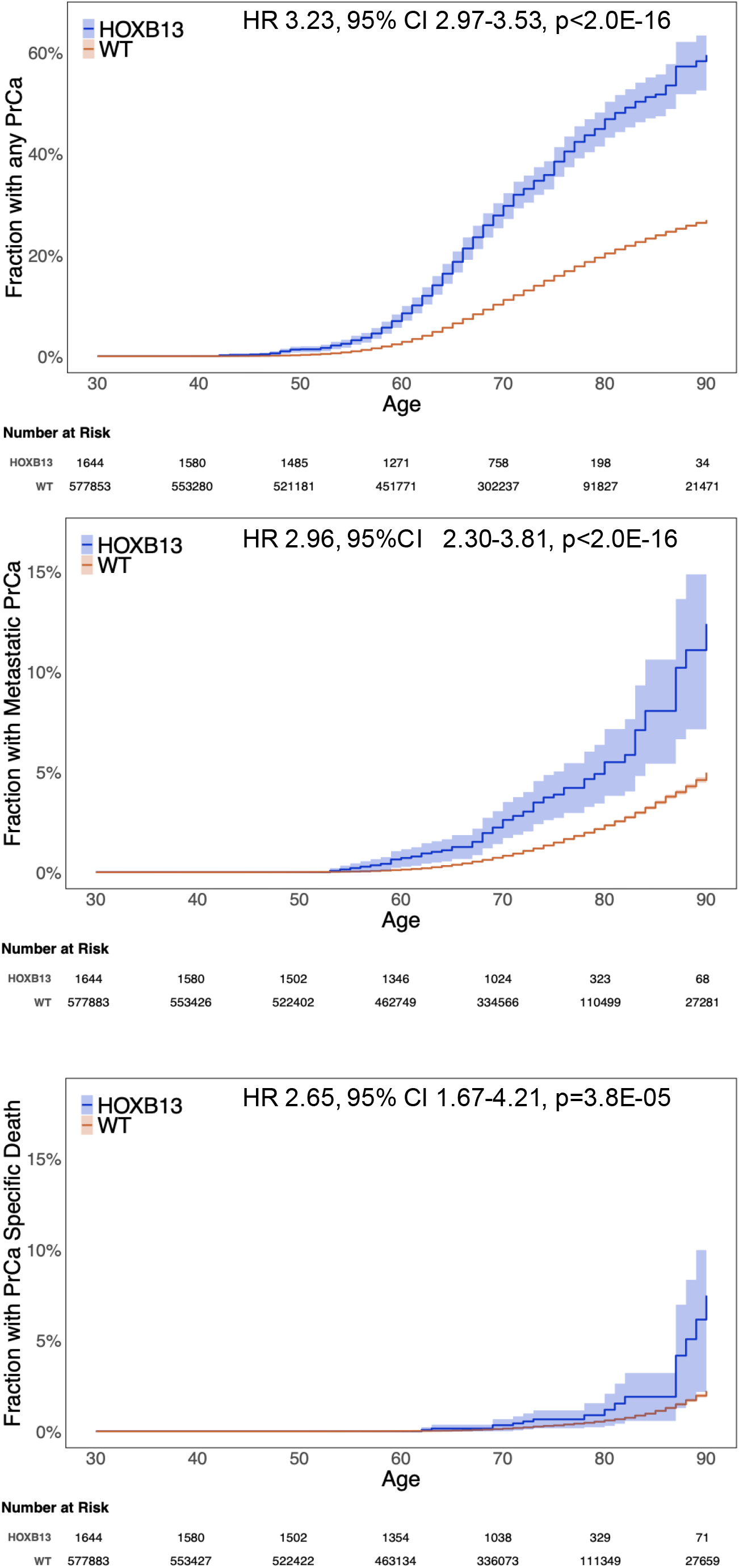
Risk of any PCa, metastatic PCa and death due to PCa in HOXB13 p.G84E heterozygotes. **A**. Cox proportional hazards model showing risk of any PCa diagnosis in *HOXB13* p.G84E heterozygous and homozygous wild-type individuals from age 25-90. Number at risk showed below the graph. **B**. Cox proportional hazards model showing risk of metastatic PCa diagnosis in *HOXB13* p.G84E heterozygous and homozygous wild-type individuals from age 25-90. Number at risk showed below the graph. **C**. Cox proportional hazards model showing risk of PCa related death in *HOXB13* p.G84E heterozygous and homozygous wild-type individuals from age 25-90. Number at risk showed below the graph.

At age 60, the fraction of heterozygotes with any PCa (8.5%) and metastatic PCa (0.7%) was higher than that of wild-type individuals (any PCa: 2.8%, metastatic PCa: 0.1%). At age 90, a larger fraction of heterozygotes had any (59.5%) or metastatic PCa (12.4%) than wild-type individuals (any PCa: 26.9%, metastatic PCa: 5.0%). (Figure 1A-B) The fraction of *HOXB13* p.G84E heterozygotes who died of PCa at age 80 and 90 was 1.2% and 7.5% respectively, compared to 0.6% and 2.2% of wild-type individuals (**Figure 1C**).

*HOXB13* p.G84E heterozygotes were more likely to be diagnosed with PCa at a younger age group (p=0.001), although the median (IQR) age of diagnosis was not statistically significantly lower overall (**Table 1**). The Gleason score (GS) distribution was similar between *HOXB13* p.G84E heterozygous and homozygous wild-type individuals. Of patients with PCa with GS data, the fraction with aggressive PCa was the same in *HOXB13* p.G84E heterozygous and homozygous wild-type individuals (33.7% vs 33.6%, p=1.0). Similar rates of de novo metastatic PCa (3.6% vs 2.9%, p=0.36), developed metastatic PCa (7.8% vs 8.9%, p=0.08), and developed metastatic castration resistant PCa (5.3% vs 6.0%, p=0.32) were observed in *HOXB13* p.G84E heterozygous and homozygous wild-type individuals (**Table 1**).

To validate our results, we analyzed 36,321 well-characterized MVP males who underwent a first prostate biopsy in the VA, among whom 202 (0.56%) were *HOXB13* p.G84E heterozygotes. *HOXB13* p.G84E heterozygotes were more likely to be diagnosed with PCa on first biopsy (83.2% vs 62.7%, p<0.001) compared to homozygous wild-type individuals (**Table S3**). In a multivariable logistic regression model, *HOXB13* p.G84E heterozygotes had a higher risk of PCa on first biopsy and risk of aggressive PCa (OR 2.60, 95%CI 1.94-3.52, p<0.001; OR 1.65, 95% CI 1.17-2.29, p=0.004) (**Table S4**). *HOXB13* p.G84E heterozygotes were diagnosed with PCa on first biopsy at a younger age (**Table S3**).

In this study, we report age-specific PCa risks and clinical-pathological associations in 1,660 individuals heterozygous for *HOXB13* p.G84E in 592,158 men genotyped in MVP, the largest unselected cohort of males with this variant to date. Additionally, our study is unique because heterozygote individuals were not identified based on high-risk families or clinical genetic testing indications. We find that *HOXB13* p.G84E confers a 3.2-fold increased risk of any PCa, lower than estimates from high-risk cohorts^3–6^ and a meta-analysis of 16 case-control studies^7^. These risk estimates were validated in a subset of well-characterized individuals diagnosed with PCa on first biopsy in the VA.

Furthermore, in contrast, to a 2016 Danish study with a smaller cohort^6^, our data showed that the variant was not associated with any aggressive PCa characteristics, outside of a slightly younger age of onset. Our results are in line with another study finding no relationship between *HOXB13* p.G84E status and high risk features^5^. It is likely only by virtue of having an increased risk of PCa that *HOXB13* p.G84E heterozygotes have a higher overall risk of developing metastatic disease and dying from PCa. This nuance should be discussed with men found to have *HOXB13* p.G84E to help target appropriate PCa screening and diagnosis strategies.

The limitation for our study includes the use of ICD diagnosis codes to define patients with PCa in the overall cohort; however, we validated our results in a subset of patients where detailed pathology results from biopsy were available. This cohort could be affected by differences of PCa screening and diagnosis across VA medical centers.

## Data Availability

It is not possible for the authors to directly share the individual-level data that were obtained from the MVP due to constraints stipulated in the informed consent. Anyone wishing to gain access to this data should inquire directly to MVP at. The data generated from our analyses are included in the manuscript main text, tables, and figures and online Supplementary Materials.

## Acknowledgements

See attached Authorship Responsibility, financial Disclosure and Acknowledgement Form

## References

1. Hamdy, F. C. et al. 10-Year Outcomes after Monitoring, Surgery, or Radiotherapy for Localized Prostate Cancer. N. Engl. J. Med. 375, 1415–1424 (2016).

2. Seibert, T. M. et al. Genetic Risk Prediction for Prostate Cancer: Implications for Early Detection and Prevention. Eur. Urol. 83, 241–248 (2023).

3. Ewing, C. M. et al. Germline Mutations in HOXB13 and Prostate-Cancer Risk. N. Engl. J. Med. 366, 141–149 (2012).

4. Xu, J. et al. HOXB13 is a susceptibility gene for prostate cancer: results from the International Consortium for Prostate Cancer Genetics (ICPCG). Hum. Genet. 132, 5–14 (2013).

5. Kote-Jarai, Z. et al. Prevalence of the HOXB13 G84E germline mutation in British men and correlation with prostate cancer risk, tumour characteristics and clinical outcomes. Ann. Oncol. 26, 756–761 (2015).

6. Storebjerg, T. M. et al. Prevalence of the HOXB13 G84E mutation in Danish men undergoing radical prostatectomy and its correlations with prostate cancer risk and aggressiveness. BJU Int. 118, 646–653 (2016).

7. Zhang, J. et al. Association between germline homeobox B13 (HOXB13) G84E allele and prostate cancer susceptibility: a meta-analysis and trial sequential analysis. Oncotarget 7, 67101–67110 (2016).

8. Hunter-Zinck, H. et al. Genotyping Array Design and Data Quality Control in the Million Veteran Program. Am J Hum Genet 106, 535–548 (2020).

9. Lee, K. M. et al. Genetic risk and likelihood of prostate cancer detection on first biopsy by ancestry. J. Natl. Cancer Inst. 116, 753–757 (2024).

10. Pagadala, M. S. et al. Polygenic risk of any, metastatic, and fatal prostate cancer in the Million Veteran Program. J. Natl. Cancer Inst. 115, 190–199 (2023).

