## Supplemental Methods for "Association of *HOXB13* G84E with prostate cancer among 592,158 men"

*Participants, genotyping, and clinical data sources*

We retrospectively obtained data from the MVP, an ongoing study that began enrollment in 2011 that includes genotying data for over 500,000 Veterans recruited from 63 Veterans Affairs medical centers across the United States^1^. MVP study participants provided blood samples for DNA extraction and genotyping using a custom Affymetrix Axiom biobank array of 723,305 variants^1^. Details on the quality control have been described previously^2^. Using direct genotyping data, genotypes at chr17:48728342 (GRCh38, Ref: C, Alt: T) were determined for each individual in MVP—homozygous reference (CC, n=782036), heterozygous alternate (CT, n=1710), and homozygous alternate (TT, n=1). The number of homozygous reference and heterozygous individuals stratified by harmonized ancestry and race/ethnicity (HARE)^2^ were compared by a Fisher’s exact test to Genome Aggregation Database (gnomAD) v4 data of matching genetic ancestries at this locus (**Table S1**). No significant deviation from gnomAD was seen for any of the four populations, demonstrating high quality genotyping at this locus. Clinical and pathological data were obtained for all individuals from the VA Corporate Data Warehouse (CDW) and the Prostate Cancer Data Core as previously described^3,4^. Men with PCa were defined as having two or more PCa ICD diagnosis codes^5^. We also identified a subset of males who underwent their first prostate biopsy in the VA as previously described^4^.

*Statistical Analyses*

Given that only one *HOXB13* p.G84E homozygous individual was found and PCa only affects males, all the analyses we performed compare male individuals heterozygous for *HOXB13* p.G84E to homozygous wildtype controls. Binary clinical and pathological variables were compared using chi-squared tests, and continuous variables were compared with Kruskal-Wallis tests with a p-value threshold of 0.05. We used a Cox proportional hazard model was used to assess associations of *HOXB13* p.G84E status with age of PCa diagnosis, metastatic PCa, and death from PCa^5^. In the subset of of patients undergoing their first prostate biopsy in VA, associations between *HOXB13* p.G84E status and risk of any PCa and aggressive PCa (defined as Gleason 4+3 and above) on first biopsy were measured using multivariable logistic regression models^4^.

**Refererences**

1.	Gaziano JM, Concato J, Brophy M, et al. Million Veteran Program: A mega-biobank to study genetic influences on health and disease. *J Clin Epidemiol*. 2016;70:214-223. doi:10.1016/j.jclinepi.2015.09.016

2.	Hunter-Zinck H, Shi Y, Li M, et al. Genotyping Array Design and Data Quality Control in the Million Veteran Program. *Am J Hum Genet*. 2020;106(4):535-548. doi:10.1016/j.ajhg.2020.03.004

3.	Alba PR, Gao A, Lee KM, et al. Ascertainment of Veterans With Metastatic Prostate Cancer in Electronic Health Records: Demonstrating the Case for Natural Language Processing. *JCO Clin Cancer Inform*. 2021;5:1005-1014. doi:10.1200/CCI.21.00030

4.	Lee KM, Nelson TJ, Bryant A, et al. Genetic risk and likelihood of prostate cancer detection on first biopsy by ancestry. *JNCI J Natl Cancer Inst*. 2024;116(5):753-757. doi:10.1093/jnci/djae002

5.	Pagadala MS, Lynch J, Karunamuni R, et al. Polygenic risk of any, metastatic, and fatal prostate cancer in the Million Veteran Program. *JNCI J Natl Cancer Inst*. 2023;115(2):190-199. doi:10.1093/jnci/djac199
